# P-wave Centric Ambulatory ECG Monitoring in Infants and Young Children

**DOI:** 10.1101/2020.06.18.20134999

**Authors:** Angela Romme, Hoang Nguyen

## Abstract

**Background:** P-wave centric ambulatory ECG monitoring has emerged as an important tool aiding the diagnosis of arrhythmias. However, with no specific pediatric approved ambulatory monitors, efficacy and user experience with these devices in young infants have not been established.

**Objective:** To evaluate tracings quality in children less than 10 kilograms who have been prescribed the P-wave centric Carnation Ambulatory Monitor (CAM) patch by Bardy Diagnostics Inc.

**Methods:** We performed an observational, retrospective study on patients prescribed 48-hour ambulatory Holter monitoring. We aimed to detail our experience with using the CAM patch with a patient population smaller than the recommended weight set forth by Bardy Diagnostics Inc. All patients less than 10 kg who were prescribed a 48-hour CAM patch were included in this review. Additionally, 2 different monitor locations (over the sternum and horizontal over the left axilla) were assessed to address the optimal placement in small children less than 10 kg.

**Results:** A total of 33 CAM reports from 25 patients, aged 0-15 months were included in the study. Mean patient age was 4.2 months ± 5.0 and mean weight was 5.3 kg ± 2.4. Twenty-Four percent of patients (8/33) had known congenital heart disease. Indications for monitoring included: tachyarrhythmia (15/33, 45%), bradycardia (6/33, 18%), ectopic rhythm (9/33, 27%), cardiac tumor (1/33, 1%), and prolonged QT interval (1/33, 1%). All CAM reports showed clear, identifiable P waves which were diagnostic and lead to changes in medical management for 30% of patients (i.e. medication adjustments or discharge from cardiology care). When comparing the P wave between a 12-lead ECG and the CAM patch, 77% of patients had the same or similar P wave morphology to lead aVF.

We found the recommended, upright placement over the sternum performed better than the horizontal placement over the left axilla for small infants and children less than 10 kg.

**Conclusion:** A P-wave centric Holter monitor is helpful in providing accurate diagnostics tracings even in infants and small children aiding in their clinical management.

## Introduction

Cardiac arrhythmias are commonly observed in infants and small children with an estimated incidence of 1/250 to 1/1000 within the general population. This incidence increases to 1/10 for patients with congenital heart disease.^1-2^ In patients with associated comorbidities, undiagnosed arrhythmias can contribute to hemodynamic instability and deterioration, adversely affecting outcomes.^3^ Given that most pediatric arrhythmias are paroxysmal in nature, their detection often relies on extended cardiac monitoring, specifically, ambulatory ECG monitoring.4 Accurate arrhythmia diagnosis in pediatrics is not without challenges, notably in infants due to awkward device placement and bulky designs5. Currently, there are no pediatric specific ambulatory monitors, resulting in a lack of experience and knowledge on the efficacy of these devices for this population. Cost effective and accessible ambulatory electrocardiogram (ECG) monitors have become an essential diagnostic tool used in the assessment and management of arrhythmias, with obvious indications for pediatric patients.6 A recent adult study found the Carnation Ambulatory Monitor (CAM) patch by Bardy Diagnostics Inc. (Seattle, WA) to have higher rhythm specificity over other ambulatory monitors, identifying more episodes of atrial tachycardia, atrial flutter, and non-sustained ventricular tachycardia.^7^ We planned to assess our use of the CAM patch in small pediatric patients for the quality of tracings and impact on clinical decision making.

## Methods

### Patients

Patients less than 10 kilograms who were prescribed 48-hour ambulatory monitoring with the Bardy CAM patch were included in a retrospective chart review from 25 patients resulting in 33 CAM reports, as some patients received multiple monitoring periods. The patients electronic medical record and the company’s web-based portal (BDxConnect.com) were accessed for data collection. Full disclosures were not available; however additional rhythm strips were able to be requested as needed. The study was approved by Rush University Medical Center Institutional Review Board.

### Monitor Placement

Rush University Medical Center, Department of Pediatric Cardiology was responsible for placing each CAM patch. Two different monitor placements were assessed. Most often, monitors were worn with the recorder in line with the sternum (Figure 1) for a duration of 48 hours. A total of five patients wore two monitors simultaneously, one in the traditional upright position over the sternum, and one with the recorder placed sideways over the lower chest, with the lower electrode wrapping around the left side of the patients back (Figure 2). Due to the lack of experience with CAM patches and patients less than 10 kg, we were unsure how placement would affect the diagnostic yield leading to the decision to trial two placements. All monitors were returned to BDX solutions to be read. Reports were uploaded to the portal in typical fashion. Bardy Diagnostics Inc. was intentionally left unware the recordings would be used for research purposes.

**Figure 1:**
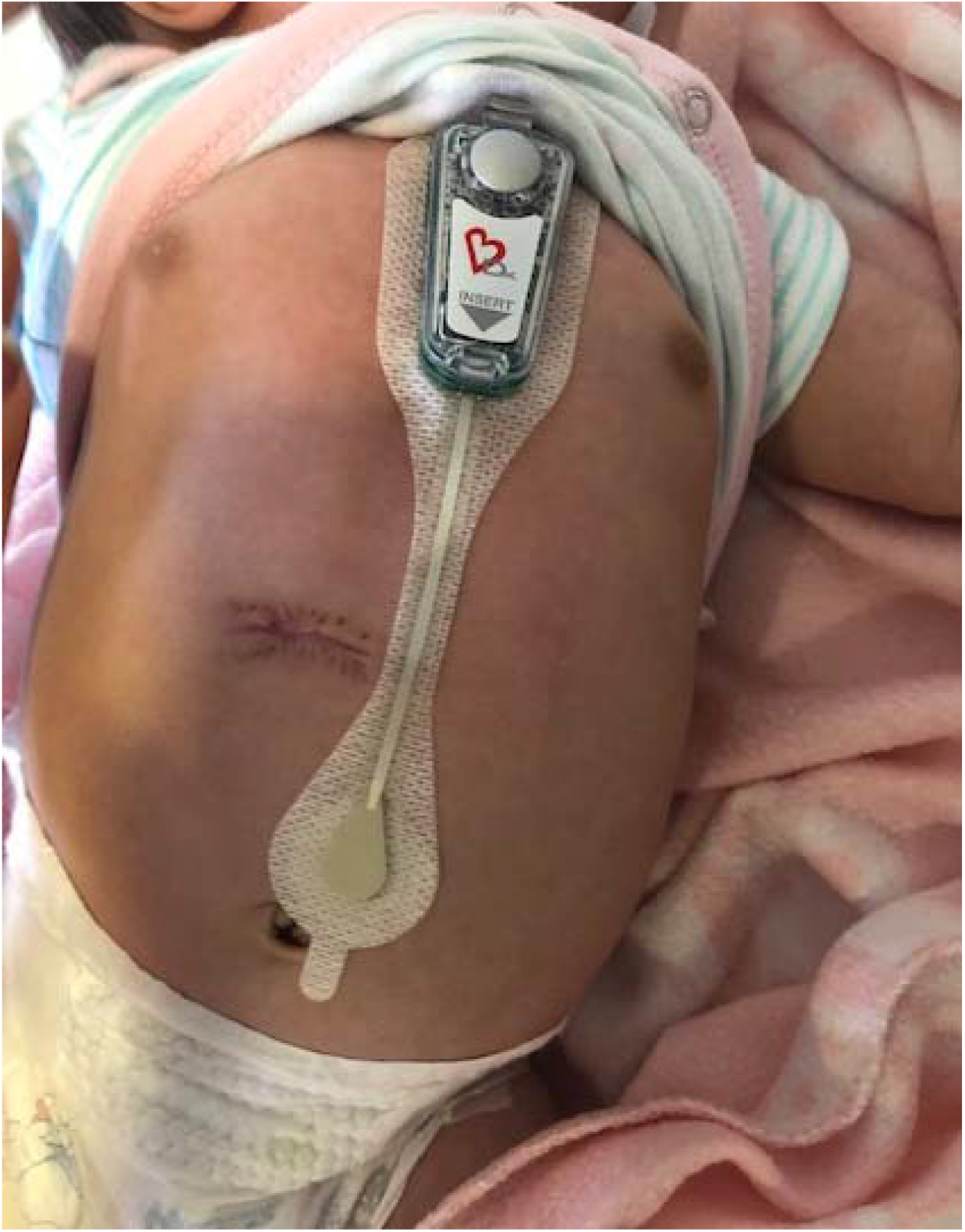
Upright device placement

**Figure 2:**
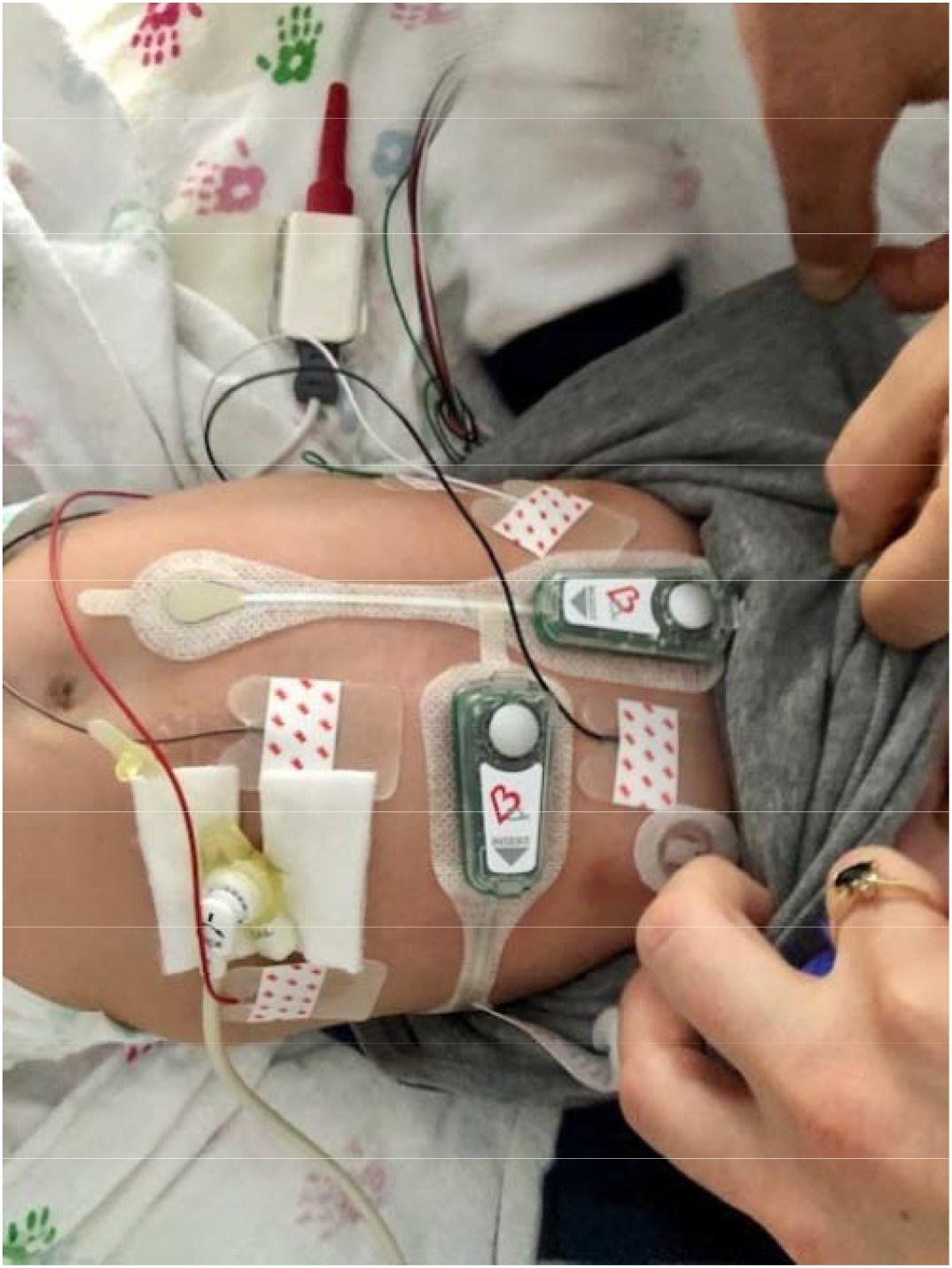
Simultaneous device placement

### Rhythm Assessment

P wave morphology and rhythm findings were compared with a standard 12-lead surface ECG performed within 24 hours of monitor placement. When applicable, continuous telemetry tracings were also used. An electrophysiologist reviewed and confirmed all findings dictated within the CAM patch report. For the five patients simultaneously wearing two monitor placements, the reports were compared against each other along with a surface ECG.

### Statistical Analysis

Summary Data is presented as percentages; Descriptive statistics are presented as a mean with range.

## Results

### Patient Demographics

Patient demographics are presented in Table 1.

**Table 1:** Demographic Data

| Demographic Data | N=33 (%) |
| --- | --- |
| Gender |  |
| *Male* | 24 (73) |
| *Female* | 9 (27) |
| Indication |  |
| *Tachyarrhythmia* | 15 (46) |
| *Bradycardia* | 6 (18) |
| *Ectopic rhythm* | 9 (27) |
| *Other* | 3 (9) |
| Clinical Setting |  |
| *Outpatient* | 18 (55) |
| *Inpatient* | 15 (45) |
| Presence of CHD | 8 (24) |
| Age (months) | 4.2 ± 5.0 |
| Weight (kg) | 5.3 ± 2.4 |

### CAM vs. 12 Lead

Thirty-One CAM reports were compared with 12-lead ECG tracings to compare P-wave characteristics. Seventy-Seven percent (24/31) of the CAM reports matched aVF morphology and axis from their corresponding 12-lead. Of note, 16% (5/31) CAM reports demonstrated increased P wave amplitude when compared to the surface ECG’s P wave. Figures 3 contains examples of P wave comparison for the same patient. A summary of the results is presented in table 2.

**Table 2:**

| Indication | CAM |  | 12 Lead ECG |  |
| --- | --- | --- | --- | --- |
|  | P wave | Interpretation | P wave | Interpretation |
| Ectopy | Upright | NSR<br>PVC <1% | Upright, normal axis | Sinus arrhythmia |
| Ectopy | Negative axis with EAR,<br>otherwise normal | Sinus Bradycardia 11%<br>PAC <1%<br>EAR <1% | Upright, normal axis | NSR, RAD |
| SVT | Upright | NSR<br>PAC <1% | Upright, normal axis | NSR, RAD |
| Bradycardia | Negative axis with EAR,<br>otherwise upright | Sinus Bradycardia<br>11%<br>PAC <1%<br>EAR | Upright, normal axis | NSR |
| NSVT | Upright | NSR<br>PAC <1% | Upright, normal axis | NSR |
| SVT | Negative axis with EAR,<br>otherwise upright | Sinus Bradycardia <1%<br>EAR <1% | Upright, normal axis | NSR |
| NSVT | Biphasic | NSVT (9 beats) | Upright, normal axis | NSR |
| EAT, Bradycardia | Upright | Sinus Bradycardia<br>2° AVB type 1<br>EAR | Upright, normal axis | NSR, RAD |
| SVT | Abnormal P waves<br>polymorphic | MAT with variable<br>conduction | Abnormal P waves<br>Polymorphic | Multifocal Atrial<br>Tachycardia |
| Tachycardia | Upright | NSR | Upright, normal axis | NSR |
| Bradycardia | Upright | NSR<br>PVC/PAC <1% | Upright, normal axis | NSR |
| Congenital Heart Block | Biphasic | CHB 99% | Upright | AV dissociation |
| Ectopy | Upright | NSR<br>EAR <1% | n/a | n/a |
| NSVT | Biphasic | NSVT (3 beats)<br>EAR | Upright, normal axis | Sinus bradycardia |
| Bradycardia | Upright | Sinus Bradycardia 15% | Upright, normal axis | NSR |

**Figure 3:**
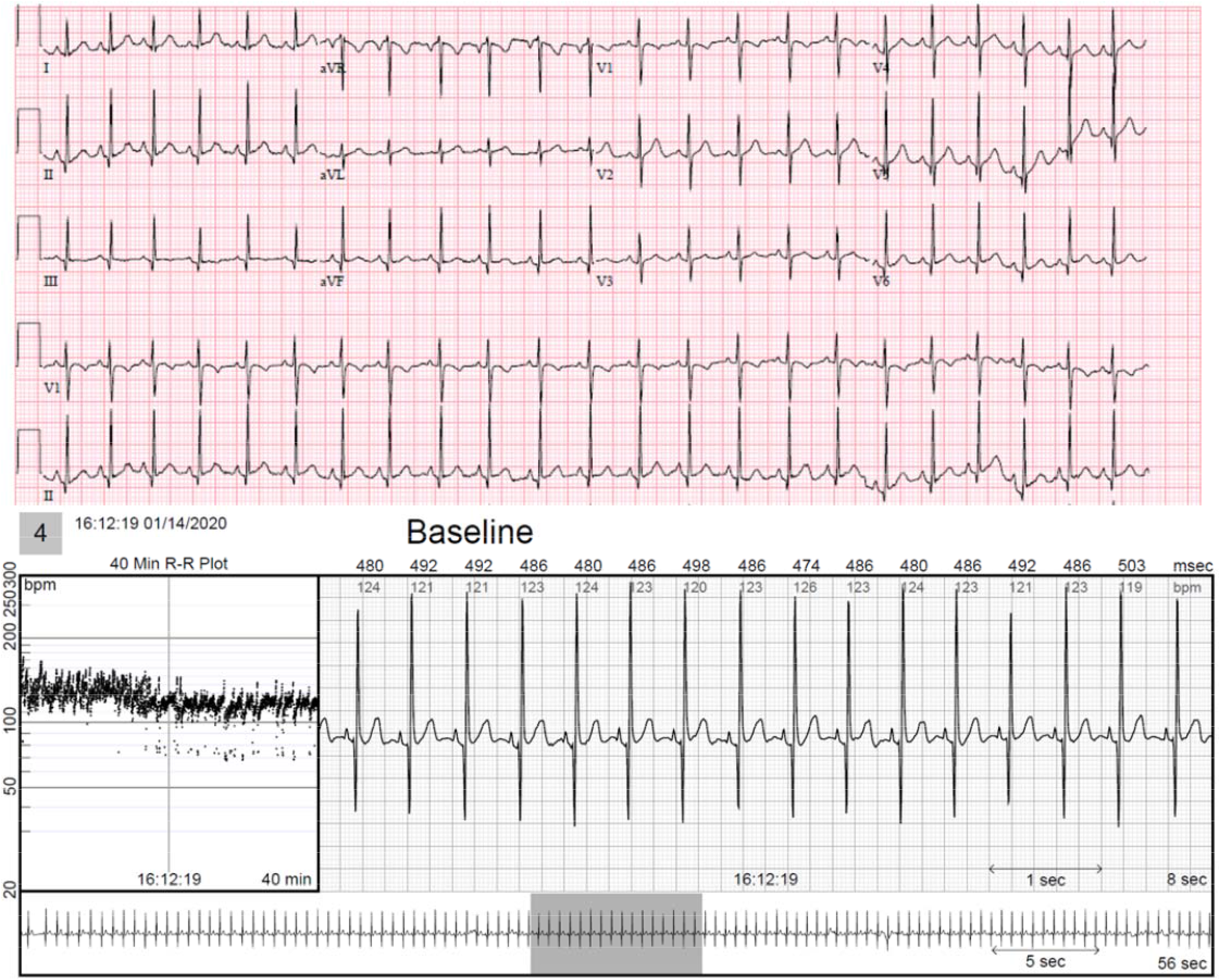
Comparis on between surface ECG and CAM strip for the same patient

### Clarity and device placement

Overall clarity was rated high by the electrophysiologist reader. P waves were identifiable in all 33 reports, with 100% interpretability for reports extracted from the recommended upright position. Five patients wore two monitors simultaneously to assess the best device orientation for patients under 10 kg (see Figure 2). The first monitor was placed according to the recommendation from Bardy, and the second monitor with the recorder placed horizontally across the lower chest with the bottom electrode extending to the patients left back. None of the horizontally placed monitors resulted in clear tracings. There was increased artifact when compared to the traditionally placed monitor, and P wave characteristics that did not match the corresponding Surface ECG, lead aVF. (See Figures 4-5.

**Figure 4:**
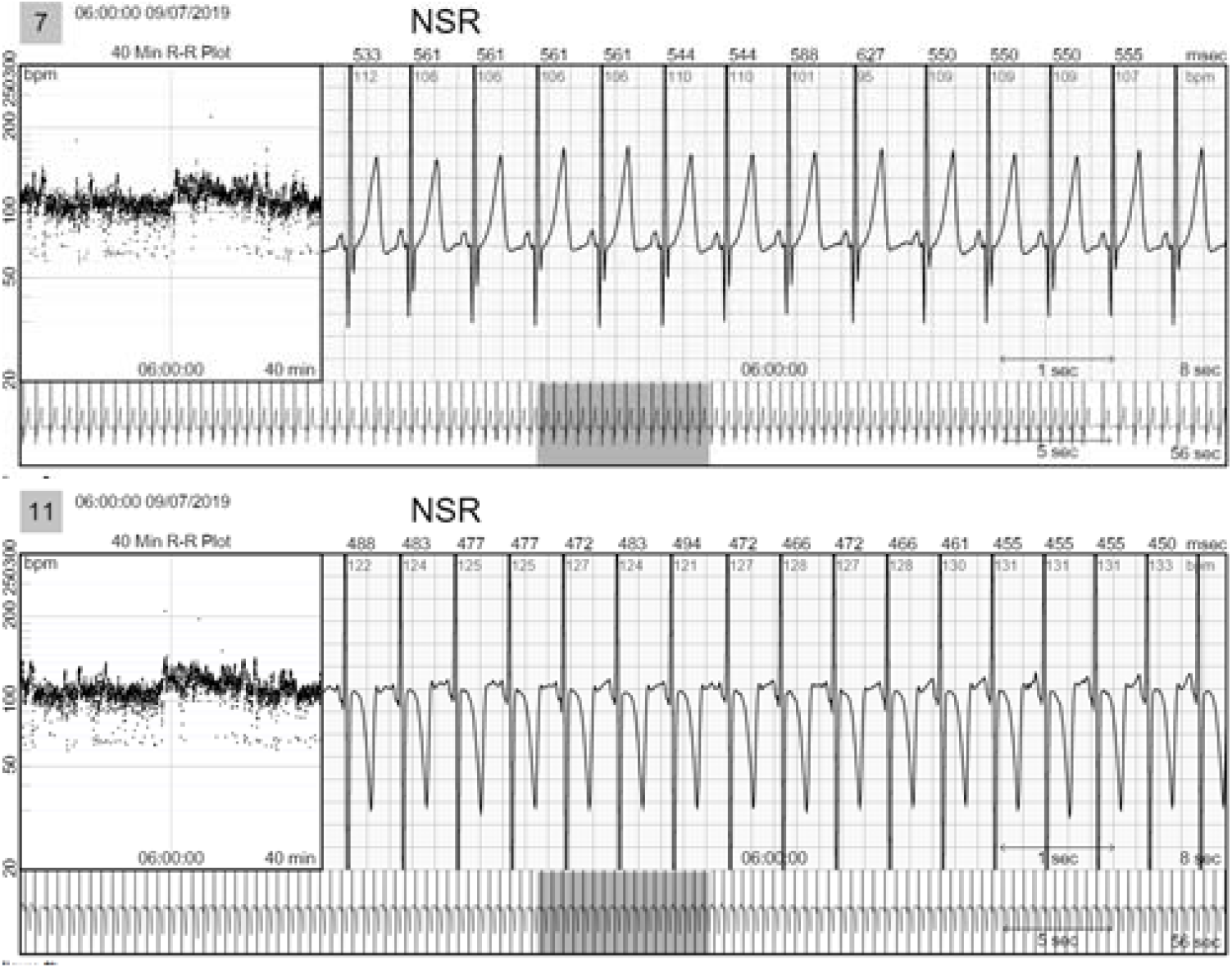
Comparison between strips taken from an upright placed monitor (above) and a horizontally placed monitor (below).

**Figure 5:**
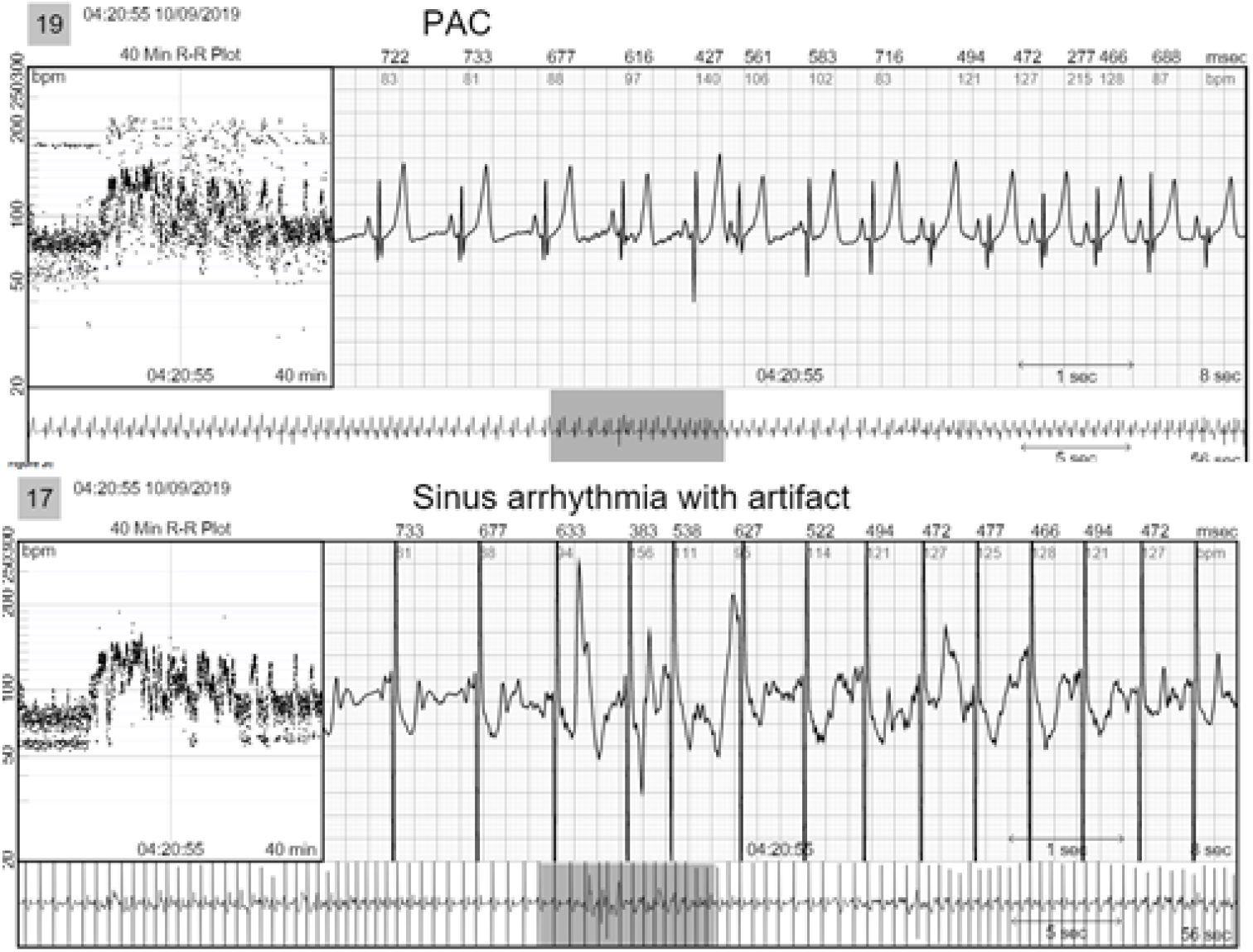
The horizontally placed monitor (below) contained significantly more artifact and did not capture the premature atrial beat that was captured from the upright monitor (above). These strips were taken at the exact same time (4:20:55)

### Tachyarrhythmia

Forty-five percent (15/33) of our patients were referred for ambulatory ECG monitoring due to concerns of tachyarrhythmia. Supraventricular tachycardia (SVT) was the most common indication for monitoring, followed by atypical atrial flutter and ectopic atrial tachycardia. A 25-day old newborn admitted to our Neonatal Intensive Care Unit (NICU) was referred a CAM patch after developing an accelerated rhythm overnight. A standard 12-lead ECG was ordered and reported an ‘Undetermined Rhythm with T wave Inversion in the Inferior Leads’ (Figure 6). The differential diagnoses included: ectopic atrial tachycardia and atrial fibrillation/flutter. A CAM patch was placed and serial ECGs were taken, along with standard NICU telemetry. The CAM patch report (Figure 7) resulted in the diagnosis of multifocal atrial tachycardia with aberrancy.

**Figure 6.**
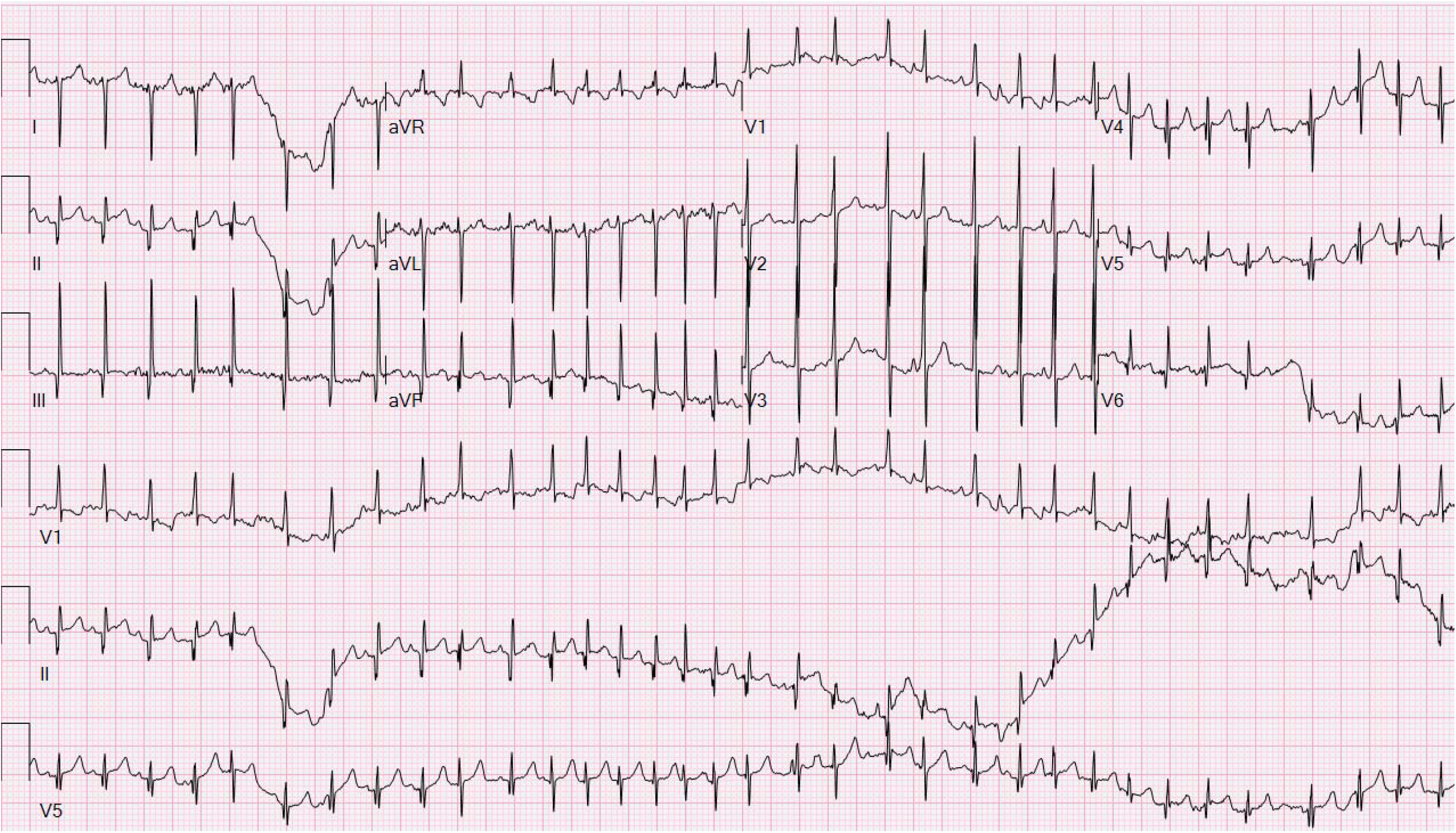
Undetermined Rhythm with T wave Inversion in the Inferior Leads

**Figure 7:**
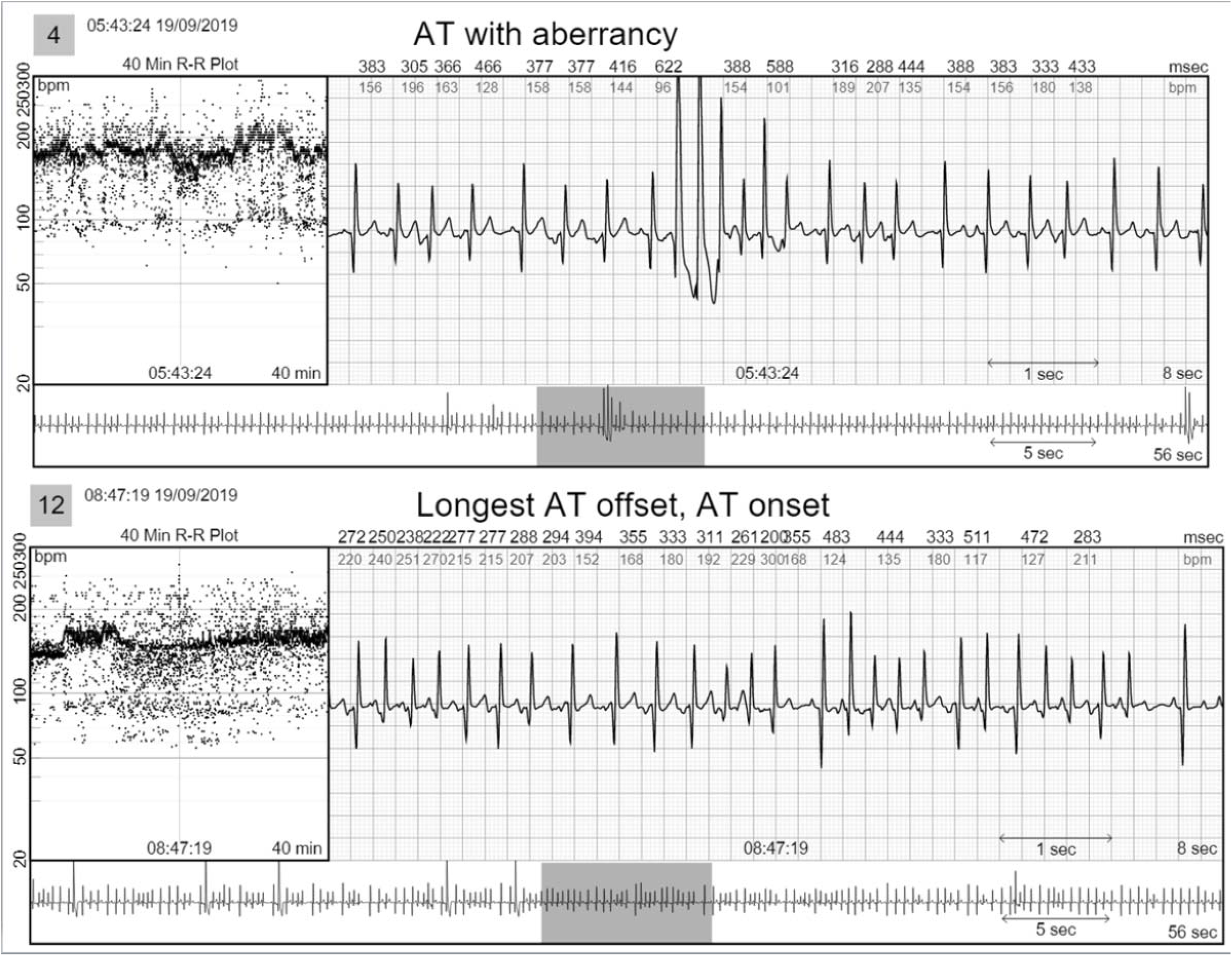
Strips from CAM patch which resulted in the appropriate diagnosis of MAT

Slightly over half (8/15, 53%) of the patients referred for tachyarrhythmia underwent a change in medical therapy, including initiation or cessation of a medication, or discharge from cardiology follow up based on CAM results.

### Bradycardia

Quantification of bradycardia according to age was demonstrated in each CAM report by a percentage (see Table 2). Additionally, heart rate variability was graphically represented to help visualize day/night heart rate trends (Figure 8).

**Figure 8:**
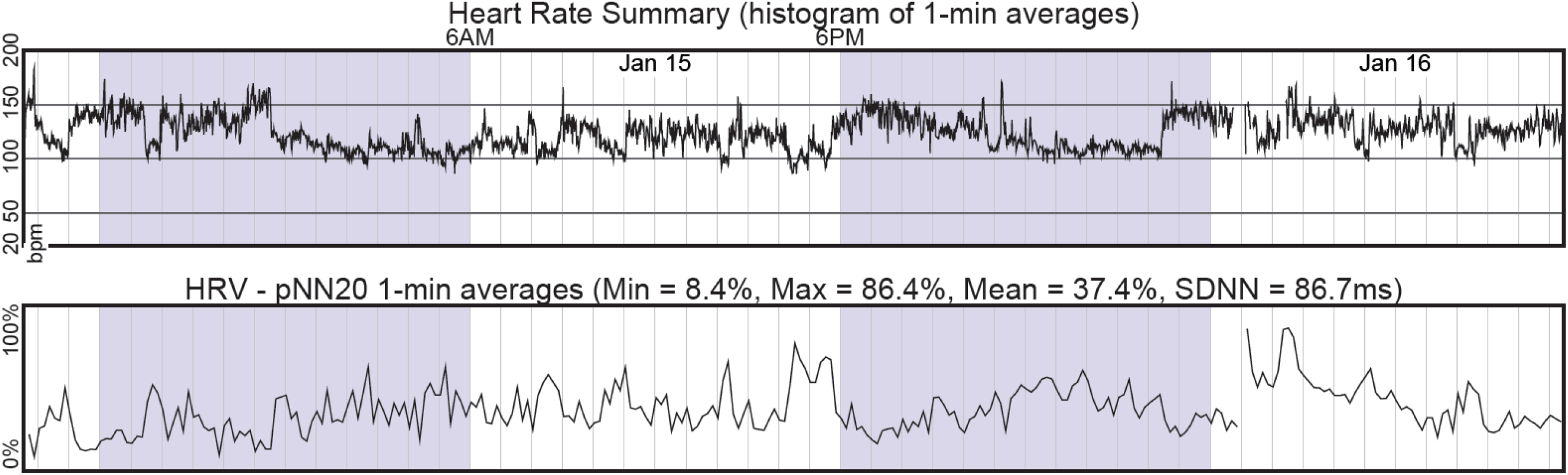
Visual representation from the CAM report for heart rate summary and heart rate variability characteristics.

### Ectopic Beats

The CAM reports clearly quantified the duration of ectopic beats or arrhythmias observed throughout the monitoring period (Figure 9). A second patient with a history of congenital heart disease admitted to the Pediatric Intensive Care Unit required an electrophysiologist assessment following intermittent ventricular tachycardia noted on telemetry. A 48-hour Bardy CAM device was placed. Comparisons between rhythm strips from telemetry and strips from the Bardy report are demonstrated in Figure 10.

**Figure 9.**
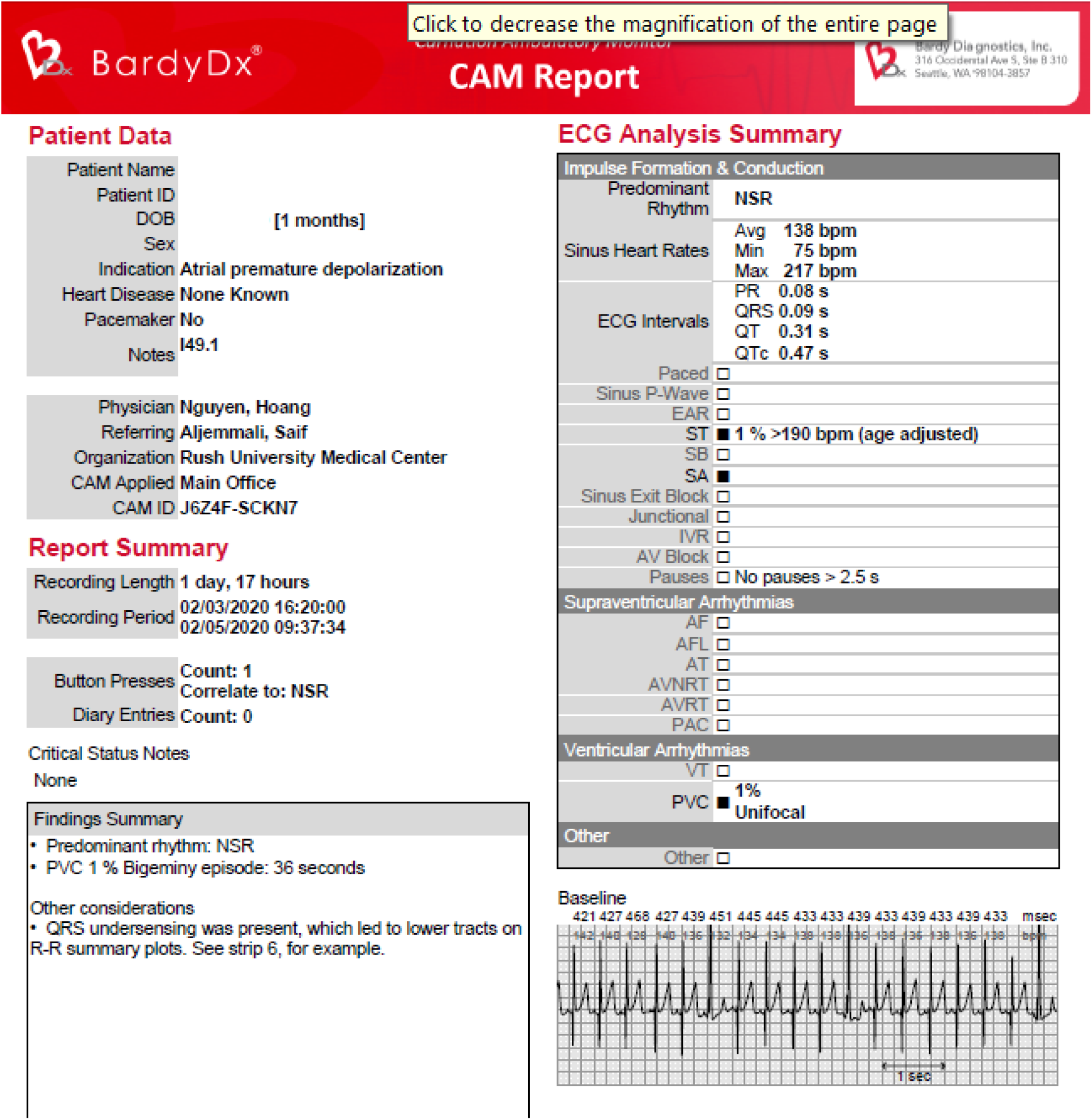
Summary page of CAM report.

**Figure 10.**
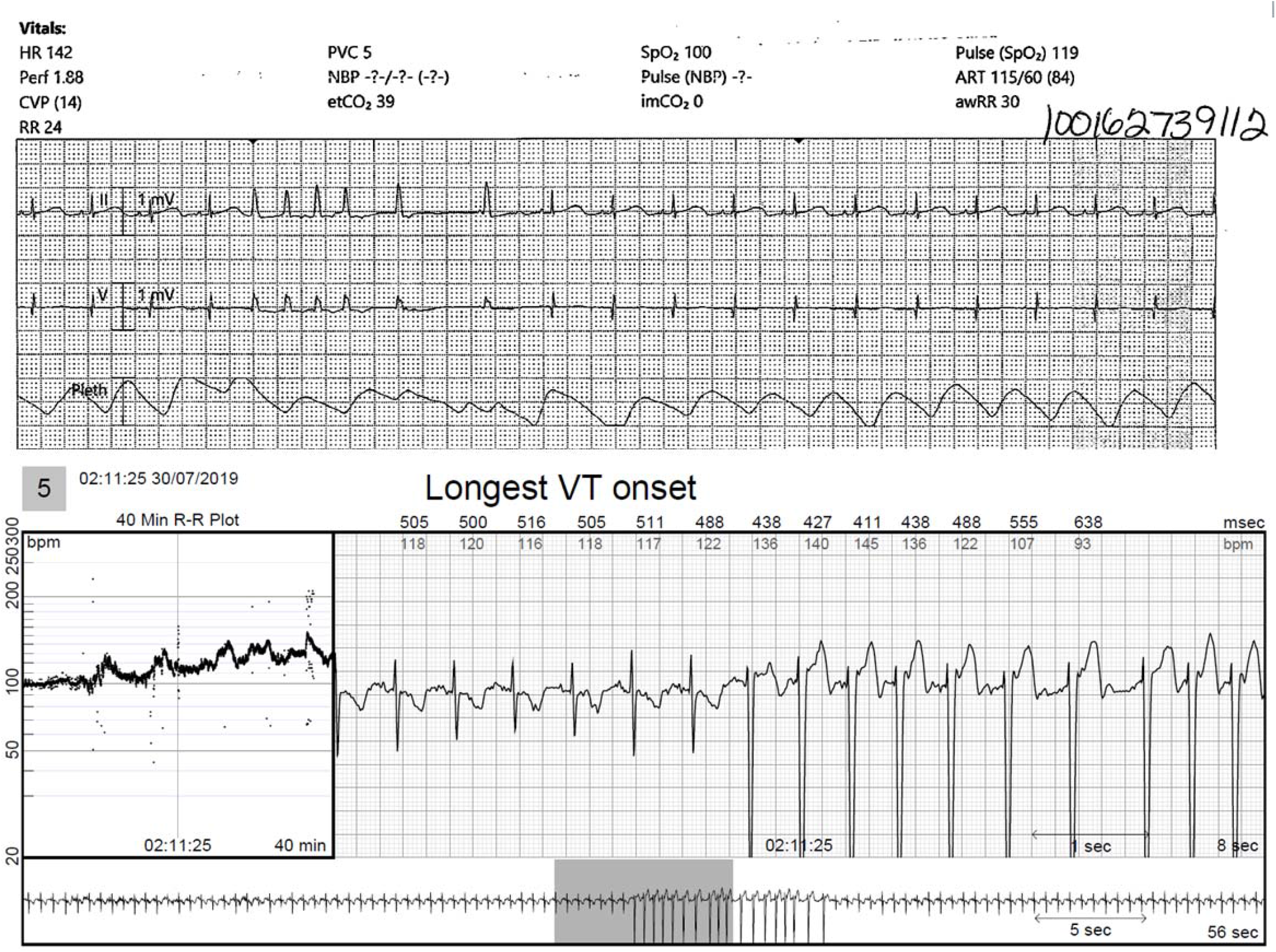

Overall, 20% (5/25) of the patients referred for ambulatory monitoring due to ectopic rhythms were able to be discharged from cardiology follow up based on the findings.

### User Experience

All patients who underwent monitoring with the CAM patch completed their studies fully. None of the studies were prematurely discontinued due to failure of monitor adhesion, or patient intolerance. None of our patients reported pressure ulcers, or skin lesions following their wear.

Our clinician experience was positive. The Bardy CAM patch is a lightweight, wireless, cutaneous single lead ECG monitor. A simple three-step set up process includes: attaching the recorder to the patch, patient skin preparation, and adhesion to the patient. Patient enrollment was completed online, through the BDxConnect portal. Once the devices were received, report generation occurred within 24-48 hours and inpatient urgent requests were accommodated.

## Discussion

Our study demonstrated the use of the CAM patch in infants and small children produced reliable data. Seventy-seven percent (24/31) of our CAM reports P waves matched the 12 lead ECG for that patient. The CAM patch utilizes a novel circuit design with compressed algorithms to process P wave signal, and placing the patch over the sternum creates a lead similar to aVF, resulting in the high sensitivity of the P-wave.^6, 7^ Traditional continuous telemetry provides a three-channel rhythm strip; however, the P wave is rarely displayed clearly.5 This is especially apparent in pediatrics, where movement artifact frequently degrades the quality of Holter recordings.^5, 7^ We were pleased to find less noise on our tracings despite the CAM patches higher P wave sensitivity. In our practice, the accurate identification of multifocal atrial tachycardia and the differentiation between atrial tachycardia with aberrancy and ventricular tachycardia resulted in optimizing medical therapy and cessation of the arrhythmia. Chu et al. found that 18% of infants suffered an adverse event after initiation of medical therapy when treating SVT with drugs other than adenosine, most commonly: hypotension, hyperkalemia, hypoglycemia, elevated liver enzymes and bradycardia.^3^ Clear P waves resulted in higher diagnostic yield and therefore the appropriate therapy, as in our pediatric patient with intermittent ventricular tachycardia. The accuracy and greater detail of the CAM helps to specify advanced arrhythmias, giving clinicians greater confidence when selecting medications, potentially decreasing the risk of an adverse event.^1^ Better rhythm assessment would be impactful in the care of congenital heart disease patients, whose hospital courses are often altered due to non-specific arrhythmia diagnoses. It is well documented that increased hospital length of stay is associated with an increased risk of adverse events.8 The Bardy CAM patch is able to identify specific rhythms that often fall under the SVT umbrella, resulting in the initiation of the appropriate therapy more rapidly.

Currently, there are no pediatric approved ambulatory monitors on the market. Guidelines set by Bardy Diagnostics Inc. for the CAM patch is for patients ten kilograms and above. We demonstrated that the P wave clarity was not compromised, and reports were diagnostic for patients less than ten kilograms. With limited experience with ambulatory monitors on young infants and children, we tested different monitor placements due to the inherent challenges of monitor orientation because of their small size. We were unsuccessful in reproducing reliable results with our horizontal placement. Our results show biphasic, inverted or unclear P waves resulting from our horizontal placement when compared to the traditionally placed monitor (See Figure 5). Importantly, ectopy noted from the traditional placement (Figure 6a) was missed and labeled as artifact in the horizontal placement (Figure 6b). This is likely due to increased movement artifact and increased the distance of the CAM recorder from the right atrium. Our findings demonstrate the recommended, upright placement of the device will still yield diagnostic results even though the device is seemingly “oversized” for these patients.

To the best of our knowledge, this study is the first to assess the efficacy of ambulatory ECG monitoring in infants and small children. Our study demonstrated the diagnostic yield of capturing arrhythmias and subsequently treating them was enhanced with the CAM patch. For cases that did not result in changes of care management, the CAM patch served a clinical purpose to discharge from cardiology follow up.

### Study Limitations

Our sample size is small; a study with increased patients may yield different results. Due to the retrospective nature of this study, we were unable to obtain ECGs and rhythm strips from the exact time periods the patients were wearing the CAM patch. Importantly, the patch cannot transmit data in real-time to the physician, which inherently delays changes to medical management based on the report findings. However, we did find the Bardy report upload to be quicker than its competitors, uploading reports within 24 to 48 hours of receiving the data. Our wear time of a 48-hour monitor is relatively short. A national registry study assessing ambulatory ECG monitors in children demonstrated that both asymptomatic and symptomatic triggered arrhythmias were detected beyond 48 hours of monitoring (44.1% and 50.4% respectively).^9,10^ Although we only used the 48-hour CAM patch, Bardy Diagnostics Inc. offers a 7 and 14 day CAM patch.

## Conclusion

The Bardy CAM patch is effective in accurately capturing rhythm assessments in infants and small children enhancing clinical decision making. However, due to the small population size, future studies are warranted.

## Data Availability

The datasets generated during and/or analysed during the current study are available from the corresponding author on reasonable request.

## Abbreviations

CAM: Carnation Ambulatory Monitor
ECG: Electrocardiogram
BPM: beats per minute
SVT: Supraventricular Tachycardia

## Statement of Conflict of Interest

The authors declare that there are no conflicts of interest.

## Notes

**Conflict of Interest:** All authors have no conflicts of interest to disclose.

**Financial Disclosure:** All authors have no financial relationships relevant to this article to disclose.

**Funding Source:** No funding was secured for this study

### Competing Interest Statement

The authors have declared no competing interest.

### Funding Statement

No funding received.

### Author Declarations

Rush University Medical Center Institutional Review Board

